# Effects of high-intensity interval training and moderate-intensity continuous training on non-motor symptoms in patients with Parkinson’s disease: a randomized pilot trial

**DOI:** 10.1101/2023.08.15.23294051

**Authors:** Ryul Kim, Seohee Choi, Nyeonju Kang, Kiwon Park, Heehyun Shin, Hanall Lee, Hyungwoo Lee, Jin-Sun Jun, Beomseok Jeon, Kyeongho Byun

**Affiliations:** Department of Neurology, Inha University Hospital, Inha University College of Medicine, Incheon, Korea; Division of Sport Science, Sport Science Institute & Health Promotion Center, Incheon National University, Incheon, Korea; Department of Mechatronics Engineering, Incheon National University, Incheon, Korea; Department of Neurology, Kangnam Sacred Heart Hospital, Hallym University College of Medicine, Seoul, Korea; Department of Neurology, Seoul National University Hospital, Seoul National University College of Medicine, Seoul, Korea

**Keywords:** Parkinson’s disease, exercise, intensity, non-motor symptoms, mood

## Abstract

**Objective:** To explore whether high-intensity interval training (HIIT) and moderate-intensity continuous training (MICT) have different effects on non-motor symptoms in patients with Parkinson’s disease (PD).

**Methods:** In this 24-week, randomized, controlled, investigator–blinded pilot trial, 33 participants with PD (Hoehn and Yahr stages 1–2; aged 50–80 years) were allocated to HIIT (3 days/week, 60% maximum aerobic power for 30–50 seconds with 1 minute rest intervals), MICT (3 days/week, 50% peak oxygen consumption), or control (usual care) groups. The primary clinical outcome was a 24-week change in the Non-Motor Symptoms Scale (NMSS) score. The secondary clinical outcomes were 24-week changes in the scores of specific non-motor questionnaires covering neuropsychiatric, sleep-related, autonomic, and sensory symptoms.

**Results:** NMSS score changes did not differ significantly among the three groups (Bonferroni-adjusted *p*>0.6 in all). In the secondary clinical outcome analyses, the MICT group showed a greater improvement in Beck Depression Inventory scores (median difference, 3.5 points; 95% confidence interval [CI], 1.4 to 6.0; Bonferroni-adjusted *p*=0.016) and, to a lesser extent, Beck Anxiety Inventory scores (median difference, 2.0 points; 95% CI, 0.0 to 10.8; Bonferroni-adjusted *p*=0.054) compared with those of the control group. However, these trends were not observed in the HIIT group when compared with the control group (Bonferroni-adjusted *p*>0.9 in all). There were no significant differences in the other secondary outcomes between the groups.

**Conclusion:** This trial did not show the potential benefits of HIIT or MICT on overall non-motor symptoms in patients with PD. However, MICT may be more effective than HIIT in alleviating mood symptoms, which requires further large-scale investigation.

**Trial registration:** CRIS (cris.nih.go.kr) identifier: KCT0007130

## INTRODUCTION

Patients with Parkinson’s disease (PD) experience various non-motor symptoms that commonly occur in the early stages of the disease and become predominant as the disease progresses.^1^ The burden of non-motor symptoms is closely associated with a poor quality of life and functional impairment in patients with PD.^2^ While a number of pharmacological approaches have been suggested for managing some non-motor symptoms in patients with PD, many available medications are of limited clinical significance and have the potential to cause side effects.^3^

The benefits of engaging in regular exercise for individuals with PD have been widely recognized.^4^ With respect to non-motor symptoms, previous studies have demonstrated the efficacy of physical exercises on cognitive function and depressive and sleep-related symptoms.^5-7^ However, it is largely unknown whether exercise improves other non-motor symptoms and how such effects differ with exercise intensity and mode.

Growing evidence in populations with cardiometabolic disease supports the superiority of high-intensity interval training (HIIT) over moderate-intensity continuous training (MICT) for improving cardiorespiratory fitness.^8^ Recent studies have shown that HIIT is feasible and safe for patients with PD,^9^ but its effects on clinical outcomes, particularly non-motor symptoms, remain unclear. Thus, this trial aimed to explore the potential effects of 24 weeks of HIIT and MICT programs on a range of non-motor symptoms in patients with PD.

## METHODS

### Study design and participants

This was a single-center, randomized controlled pilot trial. The study design was approved by the Institutional Review Board of the Inha University Hospital (2022-01-030) and registered at cris.nih.go.kr (KCT0007130). Written informed consent was obtained from all enrolled patients.

Patients with PD were recruited from the Department of Neurology at the Inha University Hospital. PD was diagnosed based on the UK PD Society Brain Bank Diagnostic Criteria. Patients who met the following criteria were included: (1) aged between 50 and 80 years, (2) had a Hoehn and Yahr stage 1 or 2, (3) had the disease duration of <5 years, (4) performed less than the recommended aerobic exercise proposed by the American College of Sports Medicine (≥20 minutes of vigorous exercise ≥3 days/week or ≥30 minutes of moderate exercise ≥5 days/week), (5) were on stable dopaminergic pharmacotherapy for □3 months, (6) and had a score ≥1 in the Movement Disorder Society Unified Parkinson’s Disease Rating Scale (MDS-UPDRS) part 1. We excluded patients who (1) had neurological, orthopedic, or cardiac comorbidities that made them unfit to perform aerobic exercise, (2) had a Montreal Cognitive Assessment (MoCA) score of <18, (3) were on antipsychotics, or (4) were unavailable for >10% of the study period.

Eligible participants were randomly allocated to the HIIT, MICT, or control groups. Randomization was performed at the Inha University Hospital Medical Statistics Support Center using a list of computer-generated random numbers (blocks of size 3). The investigators were blinded to the allocation (single-blind). We aimed for 11 participants per group, assuming 10% attrition at 6 months.^10^ A formal sample size calculation was not necessary for this pilot study. The initiation of regular drug use that could affect non-motor symptoms was not allowed during the study period.

### Exercise procedures

Appendix S1 shows the detailed exercise protocols. Each exercise session consisted of 5 minutes of warm-up, 40–60 minutes of two sets of aerobic exercise with 5 minutes of calisthenics including chair squat, chair split squat, seated dorsi flexion, and standing calf raise between the sets, and 5 minutes of cool-down. Both HIIT and MICT groups underwent aerobic exercise intervention using a cycle ergometer three days per week for 24 weeks. The total duration of aerobic exercise per session increased by 10 minutes every 8 weeks. The HIIT group started with 30 seconds of aerobic exercise at 60% of maximum aerobic power with 1 minute rest intervals for 8 weeks, gradually increasing the exercise duration by 10 seconds every 8 weeks, reaching a final exercise duration of 50 seconds per bout. The MICT group performed aerobic exercise at 50% of peak oxygen consumption (VO_2_ peak). All sessions were supervised at the study site. The control group was allowed to maintain their daily lives without the exercise intervention.

### Clinical evaluation

Non-motor symptoms were comprehensively measured using the Non-Motor Symptoms Scale (NMSS) to evaluate the overall non-motor severity, the MoCA to evaluate overall cognitive performance, the Beck Depression Inventory (BDI) to evaluate depression, the Beck Anxiety Inventory (BAI) to evaluate anxiety, the Apathy Scale (AS) to evaluate apathy, the Pittsburgh Sleep Quality Index (PSQI) to evaluate sleep quality, the Epworth Sleepiness Scale (ESS) to evaluate daytime sleepiness, the REM sleep behavior disorder screening questionnaire (RBDSQ) to evaluate RBD symptoms, the Scales for Outcomes in Parkinson’s Disease-Autonomic (SCOPA-AUT) to evaluate autonomic symptoms, the Parkinson Fatigue Scale (PFS) to evaluate fatigue, and the King’s Parkinson’s disease Pain Scale (KPPS) to evaluate pain. Motor symptoms were measured using the MDS-UPDRS part 3 during the *off*-medication state. Cardiorespiratory fitness was measured using the VO_2_ peak.

### Outcomes

The feasibility outcomes included attrition and adherence rates. Adherence was determined based on exercise frequency. The primary clinical outcome was a 24-week change in the NMSS score. The secondary clinical outcomes were 24-week changes in the scores of MoCA, BDI, BAI, AS, PSQI, ESS, RBDSQ, SCOPA-AUT, PFS, KPPS, and MDS-UPDRS part 3, VO_2_ peak, and levodopa equivalent daily dose.

### Statistical analysis

The main analysis followed the per-protocol principle, and intention-to-treat analyses were conducted as sensitivity analyses by replacing missing values with the minimum and maximum values for each group. The Shapiro–Wilk test was performed to test the normality of the data. Baseline characteristics were compared using the Kruskal–Wallis or Fisher’s exact tests, as appropriate. Within-group exercise frequency was compared using the Mann– Whitney U test. We compared the primary and secondary clinical outcomes among the three groups using the nonparametric analysis of covariance with post-hoc Bonferroni corrections. Covariates included age, sex, and the baseline value of the dependent variable. Two-sided *p* values <0.05 indicated statistical significance. All analyses were performed using *R* version (R Foundation for Statistical Computing, Vienna, Austria).

## RESULTS

### Baseline characteristics of participants

We screened 48 volunteers for eligibility between March 17, 2022, and May 31, 2022, and 33 were randomized to HIIT (*n*=11), MICT (*n*=11), and control (*n*=11) groups (Appendix S2). Baseline characteristics of the per-protocol and intention-to-treat populations did not differ significantly among the three groups (Table 1 & Appendix S3).

**Table 1.** Baseline characteristics of the per-protocol population.

| Variables | HIIT | MICT | Control | *p* value |
| --- | --- | --- | --- | --- |
| Number of subjects | 9 | 10 | 11 | – |
| Age, years | 72.0 (66.0–75.5) | 61.5 (56.5–67.8) | 65.0 (64.0–77.0) | 0.111 |
| Male sex, % | 5 (56%) | 3 (30%) | 7 (64%) | 0.346 |
| Age at PD onset, years | 68.0 (62.0–74.0) | 60.0 (54.5–64.5) | 63.0 (61.0–73.0) | 0.117 |
| PD duration, years | 2.0 (2.0–4.0) | 2.5 (1.8–3.0) | 3.0 (2.0–4.0) | 0.437 |
| MDS-UPDRS part 1 score | 4.0 (3.0–5.0) | 7.0 (3.8–10.0) | 4.0 (3.0–10.0) | 0.410 |
| MDS-UPDRS part 2 score | 4.0 (0.5–9.5) | 2.0 (0.8–2.8) | 5.0 (2.0–8.0) | 0.233 |
| MDS-UPDRS part 3 score | 22.0 (17.5–29.8) | 17.5 (13.8–27.0) | 20.0 (15.0–24.0) | 0.488 |
| NMSS score | 12.0 (10.0–24.5) | 13.0 (6.8–29.0) | 17.0 (7.0–37.0) | 0.830 |
| BDI score | 7.0 (5.5–10.5) | 8.0 (3.5–10.3) | 6.0 (4.0–9.0) | 0.786 |
| BAI score | 5.0 (3.0–6.0) | 5.0 (1.8–10.3) | 6.0 (3.0–9.0) | 0.893 |
| AS score | 16.0 (8.0–21.5) | 13.0 (9.8–15.5) | 15.0 (8.0–20.0) | 0.846 |
| MoCA score | 24.0 (22.0–28.0) | 25.0 (22.8–28.0) | 23.0 (20.0–26.0) | 0.415 |
| PSQI score | 6.0 (3.0–8.0) | 6.0 (3.0–8.3) | 4.0 (2.0–11.0) | 0.735 |
| ESS score | 6.0 (4.0–10.0) | 3.5 (2.8–7.5) | 4.0 (3.0–7.0) | 0.343 |
| RBDSQ score | 1.0 (0.5–4.0) | 5.5 (1.0–9.0) | 3.0 (0.0–7.0) | 0.298 |
| SCOPA-AUT score | 8.0 (5.0–19.5) | 12.0 (4.8–15.3) | 9.0 (5.0–13.0) | 0.972 |
| PFS score | 30.0 (23.5–38.5) | 34.5 (20.8–48.3) | 41.0 (30.0–49.0) | 0.401 |
| KPPS score | 9.0 (2.5–19.0) | 3.5 (0.8–12.0) | 4.0 (1.0–6.0) | 0.381 |
| VO<sub>2</sub> peak, mL/kg per min | 20.7 (13.8–26.5) | 20.2 (17.2–21.6) | 19.7 (14.3–23.4) | 0.984 |
| LEDD, mg | 400.0 (250.0–450.0) | 437.5 (275.0–500.0) | 400.0 (250.0–437.5) | 0.776 |

Data are n (%) and the median (interquartile range).
Abbreviations: AS=Apathy Scale; BAI=Beck Anxiety Inventory; BDI=Beck Depression Inventory; ESS=Epworth Sleepiness Scale; HIIT=high-intensity interval training; KPPS=King's Parkinson's Disease Pain Scale; LEDD=levodopa equivalent daily dose; MDS-UPDRS=Movement Disorders Society Unified Parkinson's Disease Rating Scale; MICT=moderate intensity continuous training; MoCA=Montreal Cognitive Assessment; NMSS=Non-Motor Symptoms Scale; PD=Parkinson's disease; PFQ=Parkinson's Fatigue Scale; PSQI=Pittsburgh Sleep Quality Index; RBDSQ= REM Sleep Behavior Disorder Screening Questionnaire; SCOPA-AUT=Scale for Outcomes in Parkinson's disease-Autonomic; VO<sub>2</sub> peak=peak oxygen consumption.

### Feasibility outcomes

There was no significant difference in the attrition or adherence rates between the HIIT and MICT groups. The attrition rates in the HIIT and MICT groups were 18.2% and 9.1%, respectively (*p*=1.000). Two HIIT participants dropped out of the study; one because of the excessive burden of exercise and the other because of musculoskeletal problems. One MICT participant dropped out of the study because of musculoskeletal problems. The mean exercise frequency was 2.9 (95% confidence interval [CI], 2.9 to 3.0) days/week for the HIIT group and 2.8 (95% CI, 2.7 to 3.0) days/week for the MICT group (*p*=0.934) (Appendix S4). The overall adherence rate was 95.5%.

### Clinical outcomes

Table 2 summarizes the clinical outcomes of participants. The median [interquartile range] NMSS score changes from baseline at study end were -4.0 [-6.5 to 6.0], -8.0 [-10.3 to 0.3], and -2.0 [-19.0 to 14.0] in the HIIT, MICT, and control groups, respectively. No significant differences were found among the groups (Bonferroni-adjusted *p*>0.6 in all). In the secondary clinical outcome analyses, the median [interquartile range] BDI score changes in the MICT group was -4.5 [-5.5 to -1.8] compared to -1.0 [-2.0 to 1.0] in the control group, resulting in a between-group adjusted median difference of 3.5 points (95% CI, 1.4 to 6.0 points; Bonferroni-adjusted *p*=0.016) in favor of MICT. Similarly, the improvement in BAI scores tended to be greater in the MICT group than in the control group, although the difference was not statistically significant (median difference, 2.0 points; 95% CI, 0.0 to 10.8 points; Bonferroni-adjusted *p*=0.054). However, these trends were not observed in the HIIT group when compared with the control group. The MDS-UPDRS part 3 score increases in the *off*-medication state were significantly smaller in the HIIT and MICT groups than in the control group. Moreover, both exercise groups showed significant improvements in VO_2_ peak compared with the control group. The other secondary clinical outcomes did not differ significantly among the groups.

**Table 2.** Primary and secondary clinical outcomes (by per-protocol)

| Variables | Group |  |  | Group comparisons* |  |  |
| --- | --- | --- | --- | --- | --- | --- |
|  | HIIT | MICT | Control | HIIT vs. Control | MICT vs. Control | HIIT vs. MICT |
| **Primary outcome** |  |  |  |  |  |  |
| NMSS | -4.0 (-6.5 to 6.0) | -8.0 (-10.3 to 0.3) | -2.0 (-19.0 to 14.0) | 1.000 | 0.624 | 0.821 |
| **Secondary outcomes** |  |  |  |  |  |  |
| MDS-UPDRS part 1 score | 0.0 (-3.0 to 3.5) | -3.5 (-6.3 to -2.0) | 0.0 (-3.0 to 3.0) | 1.000 | 0.720 | 1.000 |
| MDS-UPDRS part 2 score | 0.0 (-3.5 to 1.5) | -0.5 (-2.8 to 0.0) | 0.0 (-2.0 to 4.0) | 1.000 | 0.713 | 0.591 |
| MDS-UPDRS part 3 score | 1.0 (-5.3 to 2.5) | 0.5 (-4.3 to 2.0) | 4.0 (2.0 to 9.0) | **0.036** | **0.024** | 1.000 |
| BDI score | -1.0 (-2.5 to 8.0) | -4.5 (-5.5 to -1.8) | -1.0 (-2.0 to 1.0) | 1.000 | **0.016** | 0.261 |
| BAI score | 0.0 (-4.0 to 2.0) | -2.0 (-9.5 to 0.3) | 1.0 (0.0 to 3.0) | 0.903 | **0.054** | 1.000 |
| AS score | -4.0 (-9.0 to -2.0) | -3.0 (-6.3 to 1.0) | -4.0 (-5.0 to 1.0) | 0.870 | 1.000 | 1.000 |
| MoCA score | 2.0 (-1.0 to 4.5) | 0.5 (-2.0 to 1.5) | 1.0 (0.0 to 4.0) | 0.254 | 1.000 | 0.534 |
| PSQI score | -2.0 (-4.0 to -0.5) | -2.0 (-3.5 to -0.8) | -1.0 (-4.0 to 1.0) | 1.000 | 1.000 | 1.000 |
| ESS score | -1.0 (-3.5 to 0.5) | -1.0 (-2.8 to 1.3) | 0.0 (-3.0 to 4.0) | 1.000 | 0.924 | 1.000 |
| RBDSQ score | 0.0 (0.0 to 0.5) | -1.0 (-3.3 to 0.0) | 0.0 (-2.0 to 1.0) | 1.000 | 1.000 | 0.972 |
| SCOPA-AUT score | 0.0 (-2.0 to 2.0) | -2.5 (-5.5 to 4.3) | 1.0 (-1.0 to 5.0) | 0.624 | 0.249 | 1.000 |
| PFS score | 0.0 (-9.0 to 10.0) | -7.5 (-14.0 to -0.8) | -4.0 (-13.0 to 3.0) | 1.000 | 0.831 | 1.000 |
| KPPS score | -3.0 (-10.0 to 0.5) | -1.5 (-7.0 to 2.5) | -2.0 (-4.0 to 7.0) | 1.000 | 1.000 | 1.000 |
| VO<sub>2</sub> peak, mL/kg per min | 4.3 (1.3 to 7.1) | 3.4 (1.5 to 4.2) | -2.2 (-3.4 to 1.3) | **0.006** | **0.003** | 1.000 |
| LEDD, mg | 0.0 (0.0 to 25.0) | 0.0 (0.0 to 0.0) | 0.0 (0.0 to 150.0) | 1.000 | 0.243 | 1.000 |

Data are the median (interquartile range).
Bold text indicates a *p* value of less than 0.1.
\*Post hoc Bonferroni-corrected *p* values.
Abbreviations: AS=Apathy Scale; BAI=Beck Anxiety Inventory; BDI=Beck Depression Inventory; ESS=Epworth Sleepiness Scale; HIIT=high-intensity interval training; KPPS=King's Parkinson's Disease Pain Scale; LEDD=levodopa equivalent daily dose; MDS-UPDRS=Movement Disorders Society Unified Parkinson's Disease Rating Scale; MICT=moderate intensity continuous training; MoCA=Montreal Cognitive Assessment; NMSS=Non-Motor Symptoms Scale; PD=Parkinson's disease; PFQ=Parkinson's Fatigue Scale; PSQI=Pittsburgh Sleep Quality Index; RBDSQ= REM Sleep Behavior Disorder Screening Questionnaire; SCOPA-AUT=Scale for Outcomes in Parkinson's disease-Autonomic; VO<sub>2</sub> peak=peak oxygen consumption.

We repeated the analyses by imputing missing values with the minimum and maximum values from the dataset. These missing values did not alter overall clinical outcome trends (Appendix S5 & S6).

## DISCUSSION

This pilot study demonstrated the feasibility of the applied exercise protocols, as indicated by a low attrition rate of 13.6% and a high adherence rate of 95.5%, despite the challenges of the coronavirus disease pandemic. Two musculoskeletal adverse events occurred during aerobic exercise but were not serious and expected based on previous exercise trials.^10, 11^ Of note, both HIIT and MICT showed beneficial effects on cardiorespiratory fitness and motor severity, indicating that our exercise regimens were effectively implemented under the direct supervision of exercise experts in patients with PD.

Contrary to our hypothesis, neither HIIT nor MICT showed any trend toward improvement in overall non-motor symptom severity over the 24-week trial period. These observations should be interpreted with caution because of the small sample size; however, other factors may be considered to account for the lack of exercise effects. In this study, we assessed the overall non-motor symptoms of PD using the NMSS total score as the primary clinical outcome variable. As supported by our secondary clinical outcome findings, it is important to consider that the effects of exercise can vary depending on the non-motor domains.^12^ Consequently, such distinct effects may result in only modest changes in the overall severity of non-motor symptoms in the exercise groups. Furthermore, we enrolled patients with varying degrees of non-motor symptoms, ranging from mild to severe. This clinical diversity might have contributed to the increased heterogeneity in response to exercise.

Interestingly, MICT improved depressive symptoms, and to a lesser extent, anxiety symptoms, in comparison usual care, whereas such trends were not observed when HIIT was applied. These findings do not support the popular notion that the effects of exercise are intensity-dependent.^13^ Previous studies have shown that higher-intensity exercise is more beneficial for motor symptoms^10^ and sleep quality^7^ in patients with PD. However, in a recent meta-analysis of 19 randomized controlled trials involving 1302 patients with PD, the effect size of light-to-moderate-intensity exercise was slightly higher than that of moderate-to-vigorous-intensity exercise, although both exercise intensities significantly improved depressive symptoms.^6^ Similarly, a mindfulness yoga program was found to be more effective compared to stretching and resistance training exercises in reducing depressive and anxiety symptoms in patients with PD,^14^ which may emphasize the importance of exercise type over intensity in improving mood symptoms.

While the exact mechanism underlying the potential superiority of MICT over HIIT in alleviating the mood symptoms observed in this study is not fully understood, one hypothesis is that engaging in excessive exercise beyond the physical capabilities of an individual becomes a psychological burden.^15^ In addition to the difference in exercise intensity, participants in the HIIT group were older than those in the MICT group, although the difference was not statistically significant. Accordingly, HIIT program participants might have experienced a greater exercise-related psychological burden, which may explain our findings. Moreover, differences in exercise mode (intermittent vs. continuous) may partially influence the conflicting results of mood symptoms; however, how the exercise mode affects these symptoms in patients with PD remains unknown.

This study had several limitations. First, the relatively small sample size limits the ability to detect significant group effects. Second, the lack of participant blinding might have resulted in biased results owing to the potential impact of participants’ expectations or beliefs regarding the interventions. Third, the current trial specifically targeted patients with early stages of PD, which limits the generalizability of our results to the entire PD population. Finally, no adjustments were made to the multiple secondary clinical outcome analyses, thereby increasing the possibility of false positives.

In conclusion, although this pilot trial did not show the potential effect of HIIT or MICT on overall non-motor symptoms in patients with PD, MICT may be more effective than HIIT in alleviating depressive and anxiety symptoms. Further large-scale trials are warranted to clarify the effects of HIIT and MICT on mood symptoms in patients with PD.

## Data availability

The full dataset will be available on reasonable request from any qualified investigator.

## Contributorship Statement

Conception and design of the study: RK and KB. Data acquisition, analysis, and interpretation: RK, SC, NK, KP, HS, HL, HL, and KB. Statistical analysis: RK. Drafting of the manuscript: RK. Revising the manuscript: all authors.

## Notes

**Conflict of Interest Statement** The authors declare no conflicting interests.

**Funding** This work was supported by the National Research Foundation (NRF) grant funded by the Korea government (MSIT) (No. 2021R1C1C1011822).

### Competing Interest Statement

The authors have declared no competing interest.

### Clinical Trial

cris.nih.go.kr (KCT0007130)

### Funding Statement

This work was supported by the National Research Foundation (NRF) grant funded by the Korea government (MSIT) (No. 2021R1C1C1011822).

### Author Declarations

The study design was approved by the Institutional Review Board of the Inha University Hospital (2022-01-030).

### Summary of Updates

Exercise protocol revised; Appendix S1 revised.

