## Appendix for "Effects of high-intensity interval training and moderate-intensity continuous training on non-motor symptoms in patients with Parkinson’s disease: a randomized pilot trial"

### S1. Detailed exercise protocols

| |
|---|
| **A. Warming up –full body stretching (5 minutes)** |
| **B. Aerobic exercise using a cycle ergometer – Part A (20–30 minutes)** |
| *1 to 8 weeks<br>✓HIIT: aerobic exercise at 60% of maximum aerobic power for 30 seconds with 1 minute rest intervals during 20 minutes<br>✓MICT: aerobic exercise at 50% of peak oxygen consumption during 20 minutes |
| *9 to 16 weeks<br>✓HIIT: aerobic exercise at 60% of maximum aerobic power for 40 seconds with 1 minute rest intervals during 25 minutes<br>✓MICT: aerobic exercise at 50% of peak oxygen consumption during 25 minutes |
| *17 to 24 weeks<br>✓HIIT: aerobic exercise at 60% of maximum aerobic power for 50 seconds with 1 minute rest intervals during 30 minutes<br>✓MICT: aerobic exercise at 50% of peak oxygen consumption during 30 minutes |
| **C. Calisthenics – chair squat, chair split squat, seated dorsi flexion, and standing calf raise (5 minutes)** |
| **D. Rest (5 minutes)** |
| **E. Aerobic exercise using a cycle ergometer – Part B (20–30 minutes)** |
| *1 to 8 weeks<br>✓HIIT: aerobic exercise at 60% of maximum aerobic power for 30 seconds with 1 minute rest intervals during 20 minutes<br>✓MICT: aerobic exercise at 50% of peak oxygen consumption during 20 minutes |
| *9 to 16 weeks<br>✓HIIT: aerobic exercise at 60% of maximum aerobic power for 40 seconds with 1 minute rest intervals during 25 minutes<br>✓MICT: aerobic exercise at 50% of peak oxygen consumption during 25 minutes |
| *17 to 24 weeks<br>✓HIIT: aerobic exercise at 60% of maximum aerobic power for 50 seconds with 1 minute rest intervals during 30 minutes<br>✓MICT: aerobic exercise at 50% of peak oxygen consumption during 30 minutes |
| **F. Cooling down –full body stretching (5 minutes)** |

**S2. Flow diagram of patient participation**

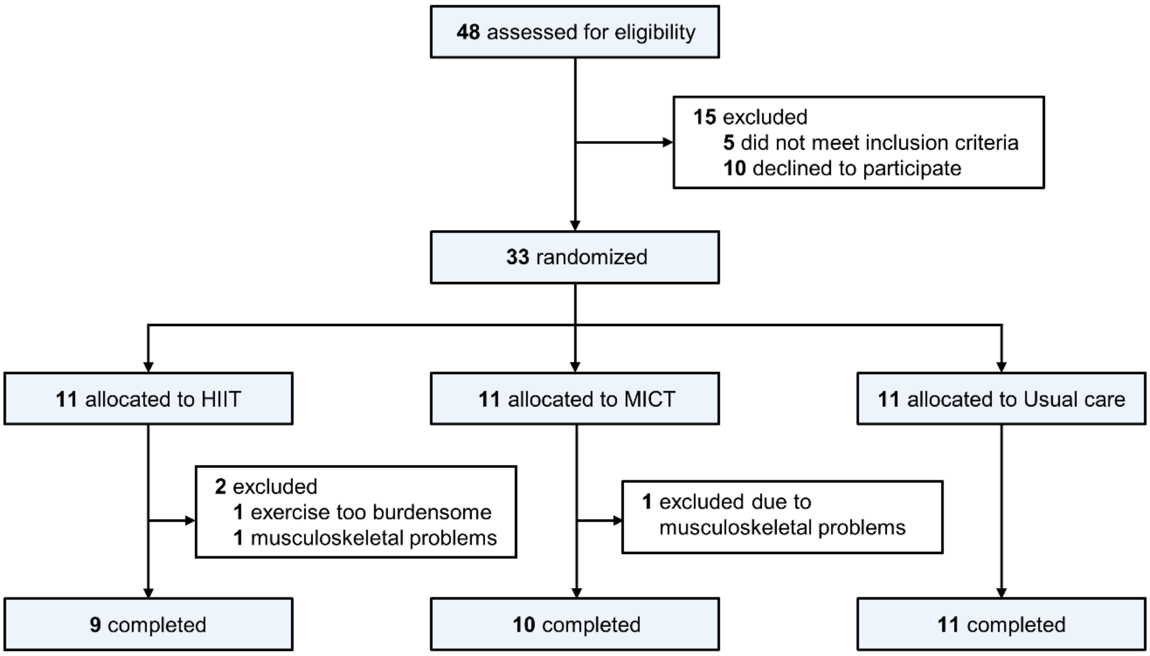


Abbreviations: HIIT=high-intensity interval training; MICT=moderate intensity continuous training.

### S3. Baseline characteristics of the intention-to-treat population

| Variables | HIIT | MICT | Control | *p* value |
|---|---|---|---|---|
| Number of subjects | 11 | 11 | 11 | – |
| Age, years | 72.0 (65.0–75.0) | 62.0 (57.0–70.0) | 65.0 (64.0–77.0) | 0.221 |
| Male sex, % | 6 (55%) | 3 (27%) | 7 (64%) | 0.307 |
| Age at PD onset, years | 68.0 (63.0–74.0) | 60.0 (55.0–66.0) | 63.0 (61.0–73.0) | 0.166 |
| PD duration, years | 2.0 (1.0–4.0) | 2.0 (2.0–3.0) | 3.0 (2.0–4.0) | 0.399 |
| MDS-UPDRS part 1 score | 4.0 (3.0–5.0) | 7.0 (4.0–10.0) | 4.0 (3.0–10.0) | 0.565 |
| MDS-UPDRS part 2 score | 4.0 (1.0–9.0) | 2.0 (1.0–5.0) | 5.0 (2.0–8.0) | 0.379 |
| MDS-UPDRS part 3 score | 22.0 (18.0–27.0) | 19.0 (14.0–27.0) | 20.0 (15.0–24.0) | 0.452 |
| NMSS score | 12.0 (10.0–24.0) | 14.0 (7.0–28.0) | 17.0 (7.0–37.0) | 0.910 |
| BDI score | 7.0 (5.0–12.0) | 7.0 (4.0–10.0) | 6.0 (4.0–9.0) | 0.710 |
| BAI score | 5.0 (4.0–8.0) | 7.0 (2.0–11.0) | 6.0 (3.0–9.0) | 0.993 |
| AS score | 15.0 (8.0–19.0) | 13.0 (10.0–16.0) | 15.0 (8.0–20.0) | 0.995 |
| MoCA score | 23.0 (21.0–28.0) | 25.0 (22.0–28.0) | 23.0 (20.0–26.0) | 0.707 |
| PSQI score | 6.0 (3.0–9.0) | 6.0 (3.0–8.0) | 4.0 (2.0–11.0) | 0.585 |
| ESS score | 6.0 (4.0–9.0) | 3.0 (2.0–7.0) | 4.0 (3.0–7.0) | 0.325 |
| RBDSQ score | 1.0 (0.0–4.0) | 6.0 (1.0–9.0) | 3.0 (0.0–7.0) | 0.212 |
| SCOPA-AUT score | 8.0 (6.0–17.0) | 12.0 (5.0–16.0) | 9.0 (5.0–13.0) | 0.913 |
| PFS score | 30.0 (19.0–42.0) | 31.0 (20.0–48.0) | 41.0 (30.0–49.0) | 0.345 |
| KPPS score | 9.0 (1.0–17.0) | 3.0 (0.0–11.0) | 4.0 (1.0–6.0) | 0.428 |
| $VO_2$ peak, mL/kg per min | 20.7 (13.8–25.1) | 20.1 (16.4–21.3) | 19.7 (14.3–23.4) | 0.936 |
| LEDD, mg | 350.0 (200.0–450.0) | 400.0 (300.0–500.0) | 400.0 (250.0–437.5) | 0.621 |

Data are n (%) and the median (interquartile range).

Abbreviations: AS=Apathy Scale; BAI=Beck Anxiety Inventory; BDI=Beck Depression Inventory; ESS=Epworth Sleepiness Scale; HIIT=high-intensity interval training; KPPS=King's Parkinson's Disease Pain Scale; LEDD=levodopa equivalent daily dose; MDS-UPDRS=Movement Disorders Society Unified Parkinson's Disease Rating Scale; MICT=moderate intensity continuous training; MoCA=Montreal Cognitive Assessment; NMSS=Non-Motor Symptoms Scale; PD=Parkinson's disease; PFQ=Parkinson's Fatigue Scale; PSQI=Pittsburgh Sleep Quality Index; RBDSQ= REM Sleep Behavior Disorder Screening Questionnaire; SCOPA-AUT=Scale for Outcomes in Parkinson's disease-Autonomic; $VO_2$ peak=peak oxygen consumption.

**S4. Mean days exercised for each intervention group during 24 weeks**

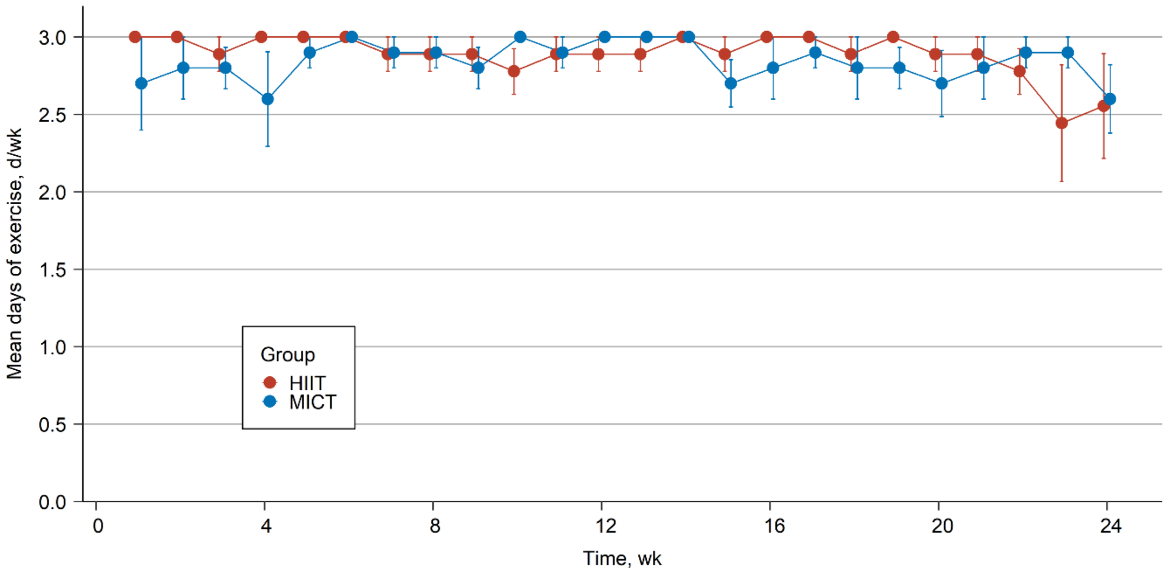


Symbols represent the means and bars are the standard errors.

#### S5. Primary and secondary clinical outcomes (by replacing missing values with the minimum values)

| | Group | | | Group comparisons* | | |
|---|---|---|---|---|---|---|
| **Variables** | **HIIT** | **MICT** | **Control** | **HIIT vs. Control** | **MICT vs. Control** | **HIIT vs. MICT** |
| **Primary outcome** | | | | | | |
| NMSS | -4.0 (-48.0 to 0.0) | -9.0 (-11.0 to -2.0) | -2.0 (-19.0 to 14.0) | 0.387 | 0.329 | 1.000 |
| **Secondary outcomes** | | | | | | |
| MDS-UPDRS part 1 score | -2.0 (-9.0 to 3.0) | -5.0 (-7.0 to -2.0) | 0.0 (-3.0 to 3.0) | 1.000 | 0.120 | 1.000 |
| MDS-UPDRS part 2 score | -1.0 (-8.0 to 1.0) | -1.0 (-5.0 to 0.0) | 0.0 (-2.0 to 4.0) | 0.979 | 0.611 | 1.000 |
| MDS-UPDRS part 3 score | -4.0 (-7.0 to 2.0) | 0.0 (-5.0 to 2.0) | 4.0 (2.0 to 9.0) | **0.010** | **0.011** | 1.000 |
| BDI score | -2.0 (-7.0 to 6.0) | -5.0 (-7.0 to -2.0) | -1.0 (-2.0 to 1.0) | 1.000 | **0.006** | 0.429 |
| BAI score | -2.0 (-11.0 to 1.0) | -3.0 (-14.0 to 0.0) | 1.0 (0.0 to 3.0) | 0.274 | **0.041** | 1.000 |
| AS score | -4.0 (-13.0 to -3.0) | -3.0 (-7.0 to 1.0) | -4.0 (-5.0 to 1.0) | 0.162 | 1.000 | 1.000 |
| MoCA score | 0.0 (-2.0 to 4.0) | 0.0 (-2.0 to 1.0) | 1.0 (0.0 to 4.0) | 1.000 | 1.000 | 1.000 |
| PSQI score | -3.0 (-6.0 to -1.0) | -2.0 (-5.0 to -1.0) | -1.0 (-4.0 to 1.0) | 1.000 | 1.000 | 1.000 |
| ESS score | -2.0 (-4.0 to 0.0) | -2.0 (-5.0 to 1.0) | 0.0 (-3.0 to 4.0) | 0.798 | 0.540 | 1.000 |
| RBDSQ score | 0.0 (-3.0 to 0.0) | -1.0 (-4.0 to 0.0) | 0.0 (-2.0 to 1.0) | 1.000 | 1.000 | 0.982 |
| SCOPA-AUT score | 0.0 (-5.0 to 2.0) | -3.0 (-7.0 to 4.0) | 1.0 (-1.0 to 5.0) | 0.231 | 0.169 | 1.000 |
| PFS score | -3.0 (-17.0 to 7.0) | -10.0 (-14.0 to -1.0) | -4.0 (-13.0 to 3.0) | 1.000 | 0.579 | 1.000 |
| KPPS score | -5.0 (-24.0 to 0.0) | -2.0 (-5.0 to 2.0) | -2.0 (-4.0 to 7.0) | 0.369 | 1.000 | 0.838 |
| $VO_2$ peak, mL/kg per min | 4.2 (-1.9 to 6.5) | 3.2 (1.2 to 3.7) | -2.2 (-3.4 to 1.3) | **0.015** | **0.006** | 1.000 |
| LEDD, mg | 0.0 (0.0 to 0.0) | 0.0 (0.0 to 0.0) | 0.0 (0.0 to 150.0) | 0.858 | 0.105 | 0.318 |

Data are the median (interquartile range).
Bold text indicates a *p* value of less than 0.1.
*Post hoc Bonferroni-corrected *p* values.
Abbreviations: AS=Apathy Scale; BAI=Beck Anxiety Inventory; BDI=Beck Depression Inventory; ESS=Epworth Sleepiness Scale; HIIT=high-intensity interval training; KPPS=King's Parkinson's Disease Pain Scale; LEDD=levodopa equivalent daily dose; MDS-UPDRS=Movement Disorders Society Unified Parkinson's Disease Rating Scale; MICT=moderate intensity continuous training; MoCA=Montreal Cognitive Assessment; NMSS=Non-Motor Symptoms Scale; PD=Parkinson's disease; PFQ=Parkinson's Fatigue Scale; PSQI=Pittsburgh Sleep Quality Index; RBDSQ= REM Sleep Behavior Disorder Screening Questionnaire; SCOPA-AUT=Scale for Outcomes in Parkinson's disease-Autonomic; $VO_2$ peak=peak oxygen consumption.

#### S6. Primary and secondary clinical outcomes (by replacing missing values with the maximum values)

| | Group | | | Group comparisons* | | |
|---|---|---|---|---|---|---|
| **Variables** | **HIIT** | **MICT** | **Control** | **HIIT vs. Control** | **MICT vs. Control** | **HIIT vs. MICT** |
| **Primary outcome** | | | | | | |
| NMSS | 0.0 (-6.0 to 13.0) | -7.0 (-10.0 to 7.0) | -2.0 (-19.0 to 14.0) | 1.000 | 0.922 | 1.000 |
| **Secondary outcomes** | | | | | | |
| MDS-UPDRS part 1 score | 2.0 (-3.0 to 6.0) | -2.0 (-6.0 to -2.0) | 0.0 (-3.0 to 3.0) | 1.000 | 0.572 | 1.000 |
| MDS-UPDRS part 2 score | 0.0 (-2.0 to 4.0) | 0.0 (-2.0 to 0.0) | 0.0 (-2.0 to 4.0) | 1.000 | 0.438 | 0.878 |
| MDS-UPDRS part 3 score | 1.0 (-4.5 to 4.0) | 1.0 (-4.0 to 2.0) | 4.0 (2.0 to 9.0) | 0.128 | **0.020** | 1.000 |
| BDI score | 2.0 (-2.0 to 14.0) | -4.0 (-5.0 to -1.0) | -1.0 (-2.0 to 1.0) | 1.000 | **0.045** | **0.066** |
| BAI score | 1.0 (-3.0 to 6.0) | -1.0 (-8.0 to 1.0) | 1.0 (0.0 to 3.0) | 1.000 | **0.097** | 0.864 |
| AS score | -4.0 (-7.0 to 1.0) | -2.0 (-6.0 to 1.0) | -4.0 (-5.0 to 1.0) | 0.563 | 1.000 | 1.000 |
| MoCA score | 3.0 (-0.0 to 6.0) | 1.0 (-2.0 to 3.0) | 1.0 (0.0 to 4.0) | 0.101 | 1.000 | 0.233 |
| PSQI score | -1.0 (-3.0 to 0.0) | -2.0 (-3.0 to 0.0) | -1.0 (-4.0 to 1.0) | 1.000 | 1.000 | 1.000 |
| ESS score | 0.0 (-3.0 to 3.0) | 0.0 (-2.0 to 2.0) | 0.0 (-3.0 to 4.0) | 1.000 | 0.814 | 1.000 |
| RBDSQ score | 0.0 (0.0 to 6.0) | -1.0 (-3.0 to 0.0) | 0.0 (-2.0 to 1.0) | 0.597 | 1.000 | 0.618 |
| SCOPA-AUT score | 1.0 (-2.0 to 3.0) | -2.0 (-5.0 to 5.0) | 1.0 (-1.0 to 5.0) | 1.000 | 0.283 | 1.000 |
| PFS score | 1.0 (-5.0 to 16.0) | -5.0 (-14.0 to 0.0) | -4.0 (-13.0 to 3.0) | 1.000 | 0.996 | 1.000 |
| KPPS score | -2.0 (-9.0 to 22.0) | 0.0 (-5.0 to 4.0) | -2.0 (-4.0 to 7.0) | 1.000 | 1.000 | 1.000 |
| $VO_2$ peak, mL/kg per min | 4.4 (2.6 to 7.7) | 3.5 (1.6 to 5.6) | -2.2 (-3.4 to 1.3) | **<0.001** | **0.003** | 1.000 |
| LEDD, mg | 0.0 (0.0 to 150.0) | 0.0 (0.0 to 0.0) | 0.0 (0.0 to 150.0) | 1.000 | 0.181 | 0.236 |

Data are the median (interquartile range).
Bold text indicates a *p* value of less than 0.1.
*Post hoc Bonferroni-corrected *p* values.
Abbreviations: AS=Apathy Scale; BAI=Beck Anxiety Inventory; BDI=Beck Depression Inventory; ESS=Epworth Sleepiness Scale; HIIT=high-intensity interval training; KPPS=King's Parkinson's Disease Pain Scale; LEDD=levodopa equivalent daily dose; MDS-UPDRS=Movement Disorders Society Unified Parkinson's Disease Rating Scale; MICT=moderate intensity continuous training; MoCA=Montreal Cognitive Assessment; NMSS=Non-Motor Symptoms Scale; PD=Parkinson's disease; PFQ=Parkinson's Fatigue Scale; PSQI=Pittsburgh Sleep Quality Index; RBDSQ= REM Sleep Behavior Disorder Screening Questionnaire; SCOPA-AUT=Scale for Outcomes in Parkinson's disease-Autonomic; $VO_2$ peak=peak oxygen consumption.
